# Immuno-inflammatory-related vulnerabilities and high mortality risk among patients with aggressive early-onset epithelial ovarian cancers in West Africa: A retrospective study

**DOI:** 10.1101/2023.07.31.23293423

**Authors:** Jude Ogechukwu Okoye, Tochukwu Juliet Ado-Okoye, Chiemeka Michael Emeka, George Uchenna Eleje, Immaculata Ogochukwu Uduchi, Uchechukwu Lilian Okoye

## Abstract

**Introduction:** This study evaluated systemic immune-inflammatory indices (SIII) among patients with epithelial ovarian cancer (EOC) to identify affordable markers for disease assessment and treatment monitoring. This study did not assess the rate of BRCA2 mutation and SIII in early-onset (≤ 50 years old) and late-onset (> 50 years old) EOC but also evaluated SIII in treatment outcomes.

**Methods:** This study included 100 patients diagnosed with EOC from Jan. 2016 to Dec. 2021. The neutrophil-to-lymphocyte ratio (NLR), platelet-to-lymphocyte ratio (PLR), platelets-neutrophils-to-lymphocytes ratio (PNLR), and neutrophils-to-lymphocytes platelets ratio (NLPR) were assessed and analyzed accordingly. Significance was set at p< 0.05.

**Result:** The frequency of early menarche, serous adenocarcinoma, and late-stage disease was 3.3, 1.6, and 1.4 times higher among patients with early-onset EOC compared with their late-onset counterparts (p= 0.001, 0.025, and 0.397, respectively). The frequency of BRCA2 mutation, hypertension, and diabetes was 2.5, 2.5, and 5.7 times higher among the latter than among the former (p= 0.001, 0.006, and 0.064, respectively). The pre-/post-treatment NLR and PNLR were 2.2/2.4 and 1.7/2.3 times higher among patients who died in the hospital than patients who were stable on discharge, respectively (p< 0.05). Although the pre-chemotherapy PNLR was 2.1 times higher among patients with stages I/II EOC compared with patients with stages III/IV EOC (p= 0.031), there was a 2.5 times significant decline and 1.1 times insignificant decline in pre-to-post-chemotherapy PNLR among the former and latter (p= 0.003 and 0.433, respectively). The post-treatment PNLR, PLR, and TWBC of herbal medicine-experienced patients were 5.6, 1.6, and 1.5, higher than the post-treatment values of naïve counterparts, respectively (p< 0.05).

**Conclusion:** This study revealed a high frequency of late-onset EOC but poor chemotherapy response among patients with aggressive early-onset. It suggests that NLR, PNLR, and PLR could be used to monitor disease progression and treatment outcomes.

## Introduction

Ovarian cancer is the third most common cancer among females in West Africa [1]. Evidence shows a higher mortality rate among women of African descent [2]. On average, there is a 2.1% annual increase in the countries within the West African region from 1995 to 2019 [3]. About 75% of ovarian cancer patients are diagnosed with late-stage disease [4,5], with a poor prognosis and a high risk of reoccurrence [6]. The incidence of epithelial ovarian cancers (EOC) is higher than other histologic types [3]. Patients with EOC have a shorter disease-free survival period [7]. Evidence shows that each five-year increase in age at menopause is associated with a 6% increase in ovarian cancer risk [8], possibly due to the increased risk of BRCA2 mutation, especially among patients who are older than 50 years [9,10]. Such BRCA-mutated ovarian cancers exhibit significantly higher inflammatory burden than BRCA wild type [11]. Patients living in countries with low/medium human development index bear the cost of cancer management. As a result, clinicians are searching for affordable prognostic systemic biomarkers to identify women with poor prognoses and assess chemotherapy response [12,13]. Systemic inflammation is linked to cancer initiation, progression, and metastasis [14]; it has been related to cancer mortality [15] and is employed as a useful prognostic indicator in many solid tumors [16]. Neutrophilia, an inflammatory process, promotes angiogenesis and immune suppression [17] while pre-treatment thrombocytosis favours advanced disease stage and high-grade epithelial ovarian cancer [7]. To improve the predictive potential of inflammatory markers, scientists are currently assessing the ratio of one immune cell to another. Studies have shown that neutrophil-to-lymphocyte (NLR), and platelet-to-lymphocyte ratio (PLR) could be used as prognostic biomarkers based on their potential to predict poor overall survival and unfavourable progression-free survival [18,19], and tumors stage [20,21]. To the best of our knowledge, this is the first study to assess BRCA2 mutation and the prognostic systemic immuno-inflammatory indices (SIII) in early-onset (≤ 50 years) and late-onset (> 50 years) EOC in West Africa. The study also evaluated changes in SIII in the disease stages and treatment outcomes.

## Methods

### Study Population and Ethics

From January 2017 to December 2021, 105 patients with ovarian cancer presented at the Department of Gynaecology, Nnamdi Azikiwe University Teaching Hospital (NAUTH), and private clinics in Nnewi and Onitsha, Nigeria. Patients with inadequate records, especially clinical and hematological data (*n* = 3) and non-epithelial tumours (n= 2) were excluded from the study. Finally, a total of 100 patients diagnosed with epithelial ovarian cancer were included. Patients received carboplatin and paclitaxel as platinum chemotherapy. This retrospective study was approved by the NAUTH ethics committee (NAUTH/CS/66/VOL.14/VER.3/35/2021/07). The patient’s medical records were accessed for socio-clinical demographics such as age, gender, comorbidities, and time of presentation. All analyses were performed by the ethical standards laid down in the Declaration of Helsinki.

### Sample collection and handling

Two samples, 5 ml of veinous whole blood, were collected from each patient and discharged into EDTA containers: a week before the first chemotherapy and a week before discharge. Full blood counts were carried out on the whole blood samples using a Haemo-autoanalyzer. Following ultrasound investigations, biopsy, and surgery, resected tissues were sent to the Department of Morbid Anatomy and Forensic Medicine for histological investigation. Two pathologists evaluated the tissues for evidence of malignancy based on the International Federation of Obstetrics and Gynecology (FIGO) guidelines. The total white cell count (TWBC) (10^^9^/L), neutrophil-to-lymphocyte ratio (NLR), platelet-to-lymphocyte ratio (PLR), platelets-neutrophils to lymphocytes ratio (PNLR; [Platelet count x Neutrophil count]/Lymphocyte count), and neutrophils-to-lymphocytes-platelets ratio (NLPR; [Neutrophil count x 100]/Lymphocyte count x platelet count]) were calculated for the subgroups.

### Procedure for Immunohistochemistry

The sections were first dewaxed and hydrated. The Epitopes in sections were then retrieved. Sections were treated with peroxidase blocker and subsequently washed in phosphate Buffered Saline (PBS). The sections were treated with the primary antibody (BReast CAncer gene 2; BRCA2) for 60 minutes in a humidity chamber, washed in PBS, and treated with the secondary antibody accordingly. The slides were then washed in PBS for 2 minutes. The sections were treated with Horseradish peroxidase and washed in 2 changes of PBS. The sections were stained with 3,3′-Diaminobenzidine (1 drop in 1 ml of Substrate), washed in PBS, stained with Haematoxylin, washed in PBS, and distilled water, dehydrated, cleared, and mounted with a DPX. The Sections were then scored based on staining intensity (0, 1, 2, and 3). Scores 0 and 1 were considered negative (mutation).

### Statistical analysis

Chi-square/Fisher was used to determine the association between the socio-clinical demographics of patients 50 years and those > 50. Pearson’s correlation was used to determine the relationship between the SIII before and after the last treatment. T-test was used for comparing data of 1. patients within aged ≤ 50 years and > 50 years, 2. chemotherapy naïve and experienced patients, 3. patients who received 1-3 cycles and 4-6 cycles of chemotherapy, 4. herbal medicine experience and naïve patients, and 5. patients with and without metastatic tumours. ANOVA was used to compare data of patients who presented at ≤ 6 months and > 6 months, and patients who were stable, unstable, and dead at discharge (in-hospital death).

## Results

The mean and median ages and age range of the patients diagnosed with ovarian cancer were 55.64 ± 13.75 years, 56.0 years, and 15 to 82 years, respectively. The reduced number of diagnoses observed in the year 2019 could be due to the limited healthcare services offered during the COVID-19 pandemic (figure 1).

**Figure 1:**
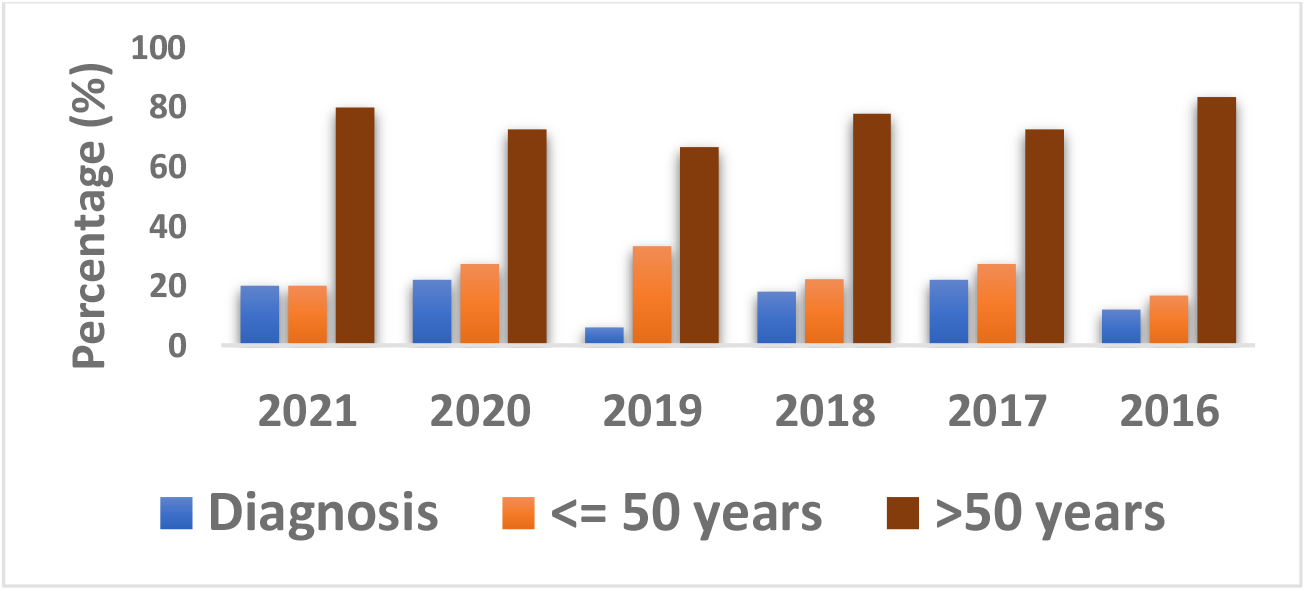
Trend of ovarian cancer cases per year. Figure 1 shows that rate of diagnosis per year was relatively the same from 2016 through 2021, except in 2019 where there was a decline in the number of cases. In 2019, the number of early-onset (≤ 50 years) cases was higher than in other years.

### Age-based socio-clinical differences

Even though the level of tertiary education was 2.7 times higher among patients ≤ 50 years old compared with their over 50 years counterparts at (p< 0.05), the unemployment rate was 3.6 times higher among the former compared with the latter at p< 0.05 (table 1). The lower employment rate among the patients aged ≤ 50 years may explain why they had a lower uptake of chemotherapy compared with their over 50 years old counterparts (p> 0.05). Although a higher percentage of the patients aged ≤ 50 years presented at the clinic within six months of symptoms manifestation compared with over 50 years counterparts (p< 0.05), the rate of in-hospital death was higher among patients aged ≤ 50 years (14/26%; 53.8%) compared with their over 50 years old counterparts (24/74; 32.4%) at p= 0.199. The median and mean survival rates were also lower among patients ≤ 50 years old (102 and 195.5 ± 67.15 years) compared with their over 50 years old counterparts (188 and 324.9 ± 71.7 years) at p= 0.317. Even though there the expression of the BRCA2 protein (figure 2) was 2.2 times higher among patients aged ≤ 50 years compared with their over 50 years counterparts (p< 0.05), the frequency of late-stage disease (stage III and IV) and serous adenocarcinoma was 1.4 and 1.6 times higher among the former than the latter at p> 0.05 and p< 0.05, respectively. The rate of early menarche (12 and 13 years) was 3.3 times higher among patients ≤ 50 years old compared with their over 50 years counterparts (p< 0.05). The frequency of hypertension was 2.5 times lower among patients aged ≤ 50 years compared with their over 50 years counterparts (p< 0.05). Although the pre-treatment systemic-inflammatory index was lower among patients aged ≤ 50 years compared with their over 50 years counterparts, the pre-treatment to post-treatment fold change TWBC, NLR, PLR, PNLR, and NLPR were relatively higher among patients who were older than 50 years (table 2).

**Table 1:** Socio-clinical characteristics of patients diagnosed with ovarian cancer.

| Variables | Total<br>N= 100 | ≤ 50 years<br>n= 26 (%) | >50 years<br>n= 74 (%) | p- value |
| --- | --- | --- | --- | --- |
| **Employment status:** |  |  |  | 0.002* |
| Civil servant | 24 | 2 (7.7) | 22 (29.7) |  |
| Unemployed/Dependant | 18 | 10 (38.5) | 8 (10.8) |  |
| Self employed | 58 | 14 (53.8) | 44 (59.5) |  |
| **Level of Education:** |  |  |  | <0.001* |
| No formal education | 9 | 1 (3.8) | 8 (10.8) |  |
| Basic education | 52 | 6 (23.1) | 46 (62.2) |  |
| Tertiary education | 39 | 19 (73.1) | 20 (27.0) |  |
| **Alcohol consumption:** |  |  |  | 0.144 |
| No | 82 | 24 (92.3) | 58 (78.4) |  |
| Yes | 18 | 2 (7.7) | 16 (21.6) |  |
| **Tobacco Use:** |  |  |  | 0.280 |
| No | 89 | 25 (96.2) | 64 (86.5) |  |
| Yes | 11 | 1 (3.8) | 10 (13.5) |  |
| **History of Hypertension:** |  |  |  | 0.006* |
| No | 52 | 20 (76.9) | 32 (43.2) |  |
| Yes | 48 | 6 (23.1) | 42 (56.8) |  |
| **Diabetes Mellitus** |  |  |  | 0.064 |
| No | 83 | 25 (96.2) | 58 (78.4) |  |
| Yes | 17 | 1 (3.8) | 16 (21.6) |  |
| **History of Herbal medicine:** |  |  |  | 0.776 |
| No | 80 | 20 (76.9) | 60 (81.1) |  |
| Yes | 20 | 6 (23.1) | 14 (18.9) |  |
| **History of fertility drug uptake:** |  |  |  | 0.726 |
| No | 88 | 24 (92.3) | 64 (86.5) |  |
| Yes | 12 | 2 (7.7) | 10 (13.5) |  |
| **Menarche:** |  |  |  | 0.001* |
| 12-13 years | 26 | 14 (53.8) | 12 (16.2) |  |
| 14-15 years | 63 | 11 (42.3) | 52 (70.3) |  |
| ≥16 years | 11 | 1 (3.9) | 10 (13.5) |  |
| **Parity:** |  |  |  | 0.474 |
| 0 | 30 | 10 (38.5) | 20 (46.2) |  |
| ≤ 2 | 14 | 4 (15.4) | 10 (13.5) |  |
| ≥ 3 | 56 | 12 (46.1) | 44 (59.5) |  |
| **TSMP:** |  |  |  | 0.009* |
| ≤ 6 months | 62 | 22 (84.6) | 40 (54.1) |  |
| > 6 months | 38 | 4 (15.4) | 34 (45.9) |  |
| **Histologic type:** |  |  |  | 0.025* |
| Serous | 64 | 23 (88.5) | 41 (55.4) |  |
| Endometrioid | 21 | 2 (7.7) | 19 (25.7) |  |
| Mucinous | 10 | 1 (3.9) | 9 (12.2) |  |
| Clear-cell | 5 | 0 (0.0) | 5 (6.7) |  |
| **FIGO Staging:** |  |  |  | 0.397 |
| Stage 1 | 24 | 4 (15.4) | 20 (27.0) |  |
| Stage 2 | 28 | 6 (23.1) | 22 (29.7) |  |
| Stage 3 | 14 | 4 (15.4) | 10 (13.5) |  |
| Stage 4 | 34 | 12 (46.1) | 22 (29.7) |  |
| **BRCA2 mutation** |  |  |  | 0.001* |
| Negative | 56 | 7 (26.9) | 49 (66.2) |  |
| Positive | 44 | 19 (73.1) | 25 (33.8) |  |
| **Chemotherapy experience:** |  |  |  | 0.165 |
| No | 41 | 14 (53.8) | 27 (36.5) |  |
| Yes | 59 | 12 (46.2) | 47 (63.5) |  |

TSMP: Time of symptom manifestation to presentation. Descriptive analysis and Chi-square/Fisher's exact test. \*Significance was set at $p < 0.05$ .

**Table 2:** Comparative analysis of hematological indices between two age groups.

| Parameters | ≤ 50 years<br>n= 26 | Fold change | > 50 years<br>n= 74 | Fold change | P -<br>value |
| --- | --- | --- | --- | --- | --- |
|  | Mean ± SD | BT/AT (%) | Mean ± SD | BT/AT (%) |  |
| **Pre-TWBC (10<sup>9</sup>/L)** | 6.75 ± 1.23 |  | 7.89 ± 1.06 |  | 0.542 |
| **Post-TWBC (10<sup>9</sup>/L)** | 5.64 ± 1.11 | 1.20 (-16.4) | 5.71 ± 0.75 | 1.38 (-27.6) | 0.958 |
| *P- value* | 0.560 |  | 0.149 |  |  |
| **Pre-NLR** | 2.79 ± 0.69 |  | 3.13 ± 0.60 |  | 0.749 |
| **Post-NLR** | 2.26 ± 0.57 | 1.23 (-19.0) | 1.85 ± 0.42 | 1.69 (40.9) | 0.641 |
| *P- value* | 0.635 |  | 0.128 |  |  |
| **Pre-PLR** | 229.5 ± 64.51 |  | 234.1 ± 33.13 |  | 0.945 |
| **Post-PLR** | 177.3 ± 50.56 | 1.29 (-22.7) | 157.9 ± 19.61 | 1.48 (-32.6) | 0.665 |
| *P- value* | 0.583 |  | 0.084 |  |  |
| **Pre-PNLR** | 792.4 ± 168.9 |  | 1031.0 ± 290.2 |  | 0.608 |
| **Post-PNLR** | 421.4 ± 72.87 | 1.88 (46.8) | 552.6 ± 155.8 | 1.86 (46.4) | 0.631 |
| *P- value* | 0.089 |  | 0.160 |  |  |
| **Pre-NLPR** | 0.22 ± 0.08 |  | 0.14 ± 0.02 |  | 0.213 |
| **Post-NLPR** | 1.21 ± 0.42 | 0.18 (450) | 0.80 ± 0.16 | 0.18 (471.4) | 0.265 |
| *P- value* | 0.008* |  | < 0.001* |  |  |

Statistics: T-test. \*Significance was set at p < 0.05.

**Figure 2:**
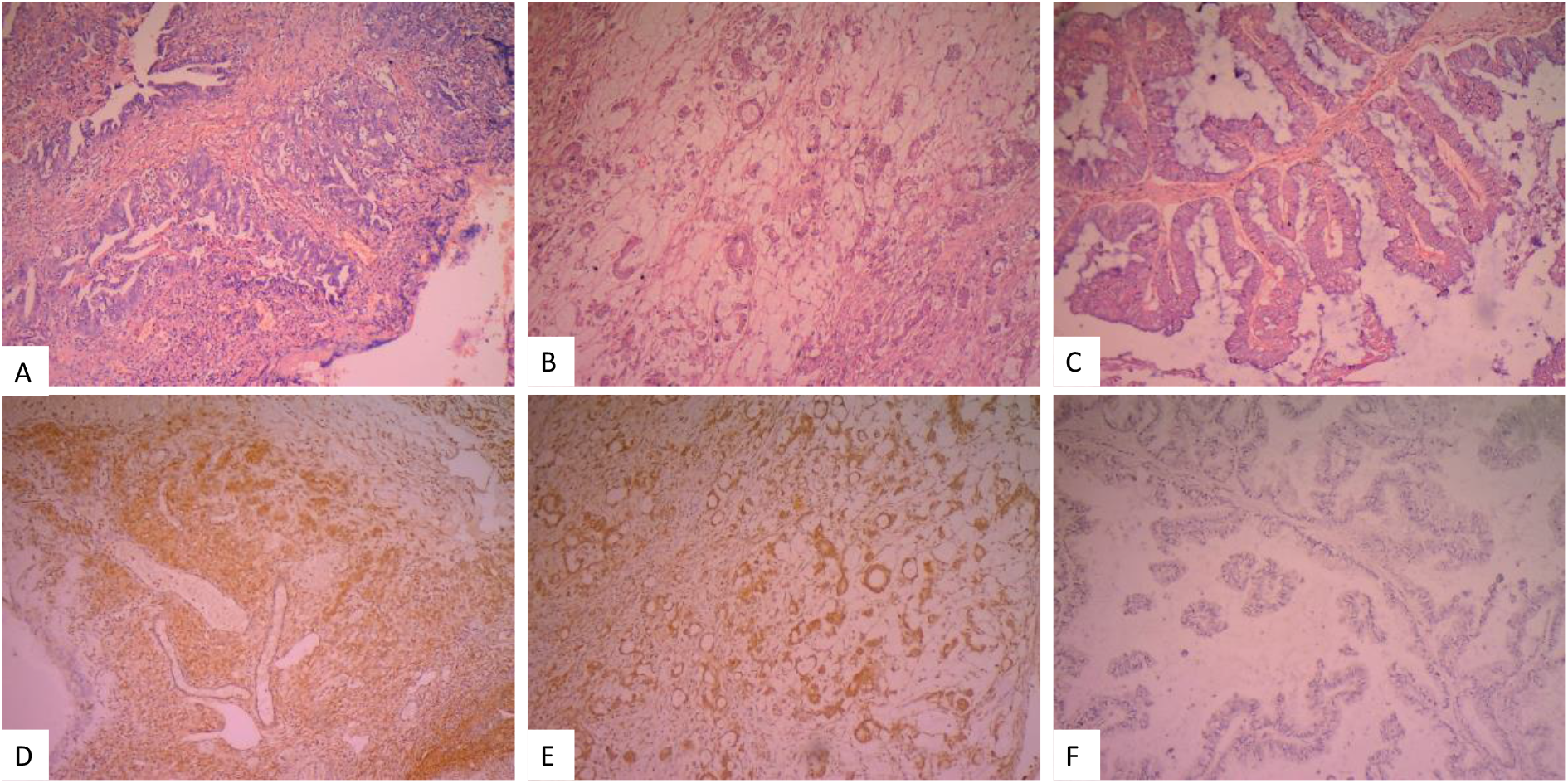
Sections of invasive ovarian cancer. Sections in figures 1A-B were stained by the H&E technique while sections in figures 1D-F were stained by the immunohistochemical technique. Sections 1D and 1E are positive for BRCA2 protein while section 1F is negative for BRCA2 protein.

### Assessment of presentation time and condition on discharge

The pre-treatment TWBC and NLPR were 1.7 and 2.0 times higher and lower among patients who presented after six months of symptom manifestation, respectively compared with patients who presented within six months of symptom manifestation at p< 0.05 (table 3). The pre-/post-treatment NLR and PNLR were 2.2/2.4 and 1.7/2.3 times higher among patients who died in the hospital, respectively compared with patients who were stable on discharge (p< 0.05). Furthermore, the fold change of pre-treatment-to-post-treatment TWBC, NLR, and PNLR was lower among patients who died in the hospital compared with patients who were stable on discharge. Conversely, the pre-treatment-to-post-treatment fold change of PLR and NLPR were higher among patients who died in the hospital compared with patients who were stable on discharge. However, the post-treatment PLR and PNLR among patients who died in the hospital were 1.4 and 1.9 times higher than the post-treatment values recorded among patients who were stable on discharge at p> 0.05, respectively (table 3). More so, patients who were stable on discharge had lower inflammatory index compared with those who died in the hospital.

**Table 3:** Comparative analysis of hematological indices based on time of symptom development to presentation and condition of the patient on discharge.

| Parameters | TSMP | Fold change<br>BT/AT (%) | TSMP | Fold change<br>BT/AT (%) | p-value |
| --- | --- | --- | --- | --- | --- |
|  | ≤ 6 Months |  | >6 Months |  |  |
|  | n= 62 |  | n= 38 |  |  |
| **Pre-TWBC ($10^9/L$)** | 4.93 ± 0.29 |  | 8.60 ± 1.07 |  | 0.001* |
| **Post-TWBC ($10^9/L$)** | 4.62 ± 0.60 | 1.07 (-6.3) | 6.72 ± 1.00 | 1.28 (-21.9) | 0.074 |
| *P- value* | 0.611 |  | 0.257 |  |  |
| **Pre-NLR** | 2.92 ± 0.54 |  | 1.66 ± 0.18 |  | 0.089 |
| **Post-NLR** | 1.58 ± 0.25 | 1.85 (-45.9) | 1.52 ± 0.21 | 1.09 (-8.4) | 0.869 |
| *P- value* | 0.066 |  | 0.626 |  |  |
| **Pre-PLR** | 204.2 ± 30.71 |  | 174.0 ± 20.79 |  | 0.457 |
| **Post-PLR** | 158.7 ± 25.31 | 1.29 (-22.3) | 152.1 ± 33.13 | 1.14 (-12.6) | 0.874 |
| *P- value* | 0.307 |  | 0.561 |  |  |
| **Pre-PNLR** | 807.1 ± 156.8 |  | 518.3 ± 56.75 |  | 0.178 |
| **Post-PNLR** | 409.1 ± 62.14 | 1.97 (-49.3) | 319.8 ± 61.32 | 1.62 (-38.3) | 0.347 |
| *P- value* | 0.038* |  | 0.033* |  |  |
| **Pre-NLPR** | 0.18 ± 0.03 |  | 0.09 ± 0.01 |  | 0.046* |
| **Post-NLPR** | 0.55 ± 0.08 | 0.32 (205.6) | 0.75 ± 0.15 | 0.12 (550.0) | 0.265 |
| *P- value* | < 0.001 |  | < 0.001 |  |  |

| Parameters | Stable OD | Fold change<br>BT/AT (%) | In-Hospital death | Fold change<br>BT/AT (%) | p-value |
| --- | --- | --- | --- | --- | --- |
|  | n= 42 |  | n= 38 |  |  |
|  | Mean ± SD |  | Mean ± SD |  |  |
| **Pre-TWBC ($10^9/L$)** | 5.75 ± 0.79 |  | 6.23 ± 0.68 |  | 0.551 |
| **Post-TWBC ($10^9/L$)** | 4.88 ± 0.84 | 1.17 (-15.1) | 5.96 ± 0.95 | 1.04 (-4.3) | 0.272 |
| *P- value* | 0.191 |  | 0.826 |  |  |
| **Pre-NLR** | 1.61 ± 0.24 |  | 3.47 ± 0.66 |  | 0.007* |
| **Post-NLR** | 1.17 ± 0.16 | 1.38 (-27.3) | 2.79 ± 0.28 | 1.24 (-19.6) | <0.001* |
| *P- value* | 0.179 |  | 0.485 |  |  |
| **Pre-PLR** | 166.2 ± 22.08 |  | 297.10 ± 62.99 |  | 0.059 |
| **Post-PLR** | 142.2 ± 19.18 | 1.17 (-14.4) | 202.60 ± 45.89 | 1.47 (-31.8) | 0.164 |
| *P- value* | 0.420 |  | 0.343 |  |  |
| **Pre-PNLR** | 591.6 ± 325.3 |  | 1016.0 ± 169.4 |  | 0.044* |
| **Post-PNLR** | 325.3 ± 35.28 | 1.82 (-45.0) | 750.5 ± 193.2 | 1.35 (-26.1) | 0.013* |
| *P- value* | 0.044* |  | 0.316 |  |  |
| **Pre-NLPR** | 0.12 ± 0.02 |  | 0.18 ± 0.04 |  | 0.166 |
| **Post-NLPR** | 0.72 ± 0.20 | 0.17 (500) | 1.40 ± 0.31 | 0.13 (677.7) | 0.070 |
| *P- value* | 0.004* |  | < 0.001* |  |  |

Keys:, OD; On discharge. Statistics: T-test and ANOVA. \*Significance was set at $p < 0.05$ .

### Comparison of chemotherapy and herbal medicine outcomes

The post-treatment TWBC and PNLR were 1.6 and 1.5 times lower among chemotherapy-experienced patients compared with chemotherapy naïve patients at p< 0.05 and p> 0.05, respectively. The post-treatment PNLR, NLPR, TWBC, NLR, and PLR of the patients who received more than three courses of chemotherapy were 2.7, 2.5, 1.9, 1.7, and 1.4 times lower than the values recorded among patients who received three or fewer courses of chemotherapy at p> 0.05, p> 0.05, p< 0.05, p> 0.05, and p> 0.05, respectively. The pre-treatment PLR, PNLR, NLPR, and NLR of herbal medicine-experienced patients were 1.9, 1.7, 1.4, and 1.1 times lower compared with the pre-treatment values of their naïve counterparts at p< 0.05, p> 0.05, p>0.05, and p> 0.05, respectively. The post-treatment PNLR, PLR, TWBC, NLR, and NLPR of herbal medicine-experienced patients were 5.6, 1.6, 1.5, 1.3, and 1.2 times higher compared with the post-treatment values of their naïve counterparts at p< 0.05, p< 0.05, p< 0.05, p> 0.05, and p> 0.05, respectively.

The pre-treatment-to-post-treatment PLR and PNLR of herbal medicine-experienced patients were significantly increased (p< 0.05) while the pre-treatment-to-post-treatment PLR and PNLR significantly declined (p< 0.05) compared with their counterparts.

The pre-treatment-to-post-treatment NLPR of both herbal medicine-experienced and naïve patients significantly increased by 5.4 and 3.4 times, respectively (p< 0.05). Of note, the median survival rate of herbal medicine-experienced and naïve patients was 86 days and 202 days, respectively, and only 50% (n= 10) of herbal medicine-experienced patients were also chemotherapy-experienced. Thus, it could be inferred that though patients with a history of herbal medicine had a lower pre-treatment SIII, they had poor treatment outcomes compared with their herbal medicine-naïve participants. Although the pre-treatment PNLR was 2.1 times higher among patients with stages I and II ovarian cancer compared with patients with stages III and IV ovarian cancer (p< 0.05), there was a 2.5 times significant decline and 1.1 times insignificant decline in the pre-to-post-treatment PNLR among the former and the latter at p< 0.05 and p> 0.05, respectively (table 4).

**Table 4:** Comparative analysis of hematological indices based on chemo/herbal therapy experience and metastasis.

| Parameters | Chemotherapy |  | p-value | Courses of Chemotherapy |  | p-value |
| --- | --- | --- | --- | --- | --- | --- |
|  | Naive | Experienced |  | 1 to 3 courses | 4 to 6 courses |  |
|  | n= 41 | n= 59 |  | n= 18 | n= 41 |  |
| | Mean ± SD | Mean ± SD | | Mean ± SD | Mean ± SD | |
| **Pre-TWBC ($10^9/L$)** | 9.29 ± 1.79 | 6.46 ± 0.70 | 0.098 | 6.14 ± 0.46 | 5.62 ± 0.43 | 0.443 |
| **Post-TWBC ($10^9/L$)** | 8.30 ± 2.10 | 5.10 ± 0.54 | 0.040* | 8.04 ± 1.48 | 4.21 ± 0.40 | 0.003* |
| *P- value* | 0.787 | 0.148 |  | 0.166 | 0.026* |  |
| **Pre-NLR** | 3.58 ± 0.82 | 2.71 ± 0.58 | 0.375 | 1.79 ± 0.21 | 1.86 ± 0.38 | 0.886 |
| **Post-NLR** | 1.72 ± 0.37 | 1.99 ± 0.42 | 0.774 | 2.11 ± 0.56 | 1.26 ± 0.16 | 0.059 |
| *P- value* | 0.253 | 0.343 |  | 0.514 | 0.182 |  |
| **Pre-PLR** | 266.4 ± 63.06 | 209.0 ± 24.31 | 0.347 | 152.0 ± 11.95 | 244.9 ± 28.67 | 0.042* |
| **Post-PLR** | 168.2 ± 40.56 | 161.7 ± 21.94 | 0.899 | 178.1 ± 62.78 | 129.7 ± 14.32 | 0.269 |
| *P- value* | 0.423 | 0.159 |  | 0.628 | 0.002* |  |
| **Pre-PNLR** | 1144 ± 407.9 | 880.9 ± 245.8 | 0.573 | 454.4 ± 67.15 | 525.8 ± 78.99 | 0.523 |
| **Post-PNLR** | 718.6 ± 306.5 | 473.1 ± 126.7 | 0.424 | 929.7 ± 515.3 | 343.2 ± 52.04 | 0.108 |
| *P- value* | 0.492 | 0.143 |  | 0.339 | 0.060 |  |
| **Pre-NLPR** | 0.19 ± 0.06 | 0.15 ± 0.03 | 0.543 | 0.11 ± 0.03 | 0.12 ± 0.03 | 0.868 |
| **Post-NLPR** | 0.52 ± 0.14 | 0.99 ± 0.19 | 0.263 | 1.74 ± 0.59 | 0.71 ± 0.14 | 0.039* |
| *P- value* | 0.020* | < 0.001* |  | 0.008* | <0.001* |  |

| Parameters | Herbal medicine |  | p- value | FIGO Stages |  | p-value |
| --- | --- | --- | --- | --- | --- | --- |
|  | Naive | Experienced |  | Stages I & II | Stages III & IV |  |
|  | n= 79 | n= 21 |  | n= 52 | n= 48 |  |
| **Pre-TWBC ($10^9/L$)** | 5.63 ± 0.35 | 5.88 ± 0.88 | 0.728 | 5.60 ± 0.35 | 5.41 ± 0.38 | 0.743 |
| **Post-TWBC ($10^9/L$)** | 4.54 ± 0.32 | 7.18 ± 1.59 | 0.013* | 4.68 ± 0.61 | 5.08 ± 0.48 | 0.641 |
| *P- value* | 0.037* | 0.358 |  | 0.169 | 0.586 |  |
| **Pre-NLR** | 1.70 ± 0.18 | 1.55 ± 0.28 | 0.669 | 2.03 ± 0.27 | 2.37 ± 0.51 | 0.526 |
| **Post-NLR** | 1.46 ± 0.17 | 1.97 ± 0.51 | 0.267 | 1.31 ± 0.19 | 1.85 ± 0.25 | 0.098 |
| *P- value* | 0.344 | 0.454 |  | 0.072 | 0.380 |  |
| **Pre-PLR** | 236.7 ± 22.38 | 127.5 ± 14.40 | 0.010* | 209.0 ± 24.33 | 190.9 ± 27.54 | 0.646 |
| **Post-PLR** | 140.5 ± 15.08 | 222.5 ± 46.32 | 0.039* | 161.5 ± 28.55 | 165.0 ± 23.30 | 0.928 |
| *P- value* | 0.002* | 0.030* |  | 0.218 | 0.490 |  |
| **Pre-PNLR** | 744.1 ± 107.1 | 431.0 ± 76.52 | 0.178 | 843.1 ± 130.1 | 410.5 ± 40.26 | 0.031* |
| **Post-PNLR** | 348.8 ± 42.60 | 1950.0 ± 541.8 | <0.001* | 326.2 ± 50.76 | 369.2 ± 32.61 | 0.518 |
| *P- value* | 0.001* | 0.010* |  | 0.003* | 0.433 |  |
| **Pre-NLPR** | 0.15 ± 0.02 | 0.11 ± 0.03 | 0.303 | 0.18 ± 0.04 | 0.14 ± 0.03 | 0.504 |
| **Post-NLPR** | 0.51 ± 0.07 | 0.59 ± 0.16 | 0.598 | 0.61 ± 0.10 | 0.70 ± 0.13 | 0.232 |
| *P- value* | < 0.001* | 0.001* |  | < 0.001* | < 0.001* |  |

Statistics: T-test. \*Significance was set at $p < 0.05$ .

## Discussion

This study compared the clinical features of early-onset (≤ 50 years) and late-onset (> 50 years) EOC. It also assessed the ratio of immune cells in different timelines of disease presentation and stages, and treatment history and outcomes. The frequency of late-onset ovarian cancer in this study was 2.8 times higher than the frequency of early-onset EOC. The high mean and median ages within our cohort agree with earlier studies’ reports showing that older age and BRCA mutation increases the risk of developing ovarian cancer [8-10]. Late-onset EOC in this study is higher than in cases diagnosed from 2000 to 2013 (32.4% to 40%) in other parts of West Africa [22-24]. The reason for the variation is unknown but it could be associated timeline of diagnosis. This is underscored by the findings of Okunade et al. who earlier recorded a low frequency of late-onset EOC in western Nigeria between 2008 and 2012 (40%) and reported a higher frequency of 62% between 2010 and 2018 in the same city [23,25]. The high frequency of late-onset EOC may be due to the high frequency of diabetes mellitus within the group [26].

In this study, higher pre-and post-treatment TWBC and SIII were observed in cases of in-hospital death compared to patients who were stable on discharge. This suggests that SIII could be used for prognostication. Li et al. reported that patients with NLR > 2.72 and PLR > 219.00 were significantly associated with decreased OS and DFS [27]. This aligns well with the values recorded in cases of in-hospital death in this study. Although most of the patients with early-onset EOC had lower frequencies of BRCA2 mutation, tobacco, alcohol, and fertility drug use, they had a higher frequency of late-stage disease compared to patients with late-onset EOC. Of note, patients who presented within six months of symptom manifestation had relatively higher pre-treatment-to-post-treatment NLR, PLR, and PNLR compared to patients who presented after six months. Since most patients with early-onset EOC presented within six months of symptom manifestation yet developed late-stage EOC, it could be argued that early-onset EOCs were aggressive and had a poorer treatment outcome than late-onset EOCs. The aggressive nature of their disease may also explain why they had a lower fold change of pre-to-post-treatment TWBC, PLR, NLR, and NLPR compared to patients with late-onset EOC. The aggressiveness of their disease may be attributed to high frequencies of early menarche and serous adenocarcinoma within the group [8,28], resulting in low survival compared to patients with late-onset EOC.

In this study, the prevalence of herbal medicine use is high but lower than the prevalence (28.3% to 73.8%) reported in other parts of West Africa [29]. Due to the high cost of orthodox treatment in countries with low healthcare resources, patients resort to herbal products. Although positive results have been recorded in vitro studies that evaluated the anti-ovarian cancer properties of some herbal products [30,31], controversies regarding the in vivo health benefits of herbal therapies abound, especially in Africa. Studies have shown that medicinal plants contain pesticides, radioelements, microorganisms, and heavy metals such as Lead, arsenic, mercury, Cadmium, Chromium, Nickel, and Zinc [32,33]. Apart from the potential to induce anaemia, diarrhea, vomiting, and electrolyte imbalance, some herbal products are believed to inhibit enzymes, especially CPY isoforms, involved in drug metabolism, resulting in increased levels and toxicity [34]. The latter could be the explanation for the higher post-treatment TWBC and SIII and lower median survival rate among herbal medicine-experienced patients compared with their herbal medicine-naïve counterparts. However, the contents of herbal medicine used by the patients in this study were not assessed might constitute a limitation. Another limitation is the small population size of the early-onset EOC in this study. This could be the reason for the insignificant differences in SIII values observed between early-onset and late-onset.

## Conclusion

This study unveils compelling findings regarding patients diagnosed with EOC in West Africa. It reveals a high frequency of late-onset EOC. It suggests that age at menarche and histologic type drive the aggressiveness of early-onset EOC and determine treatment outcome. It also reveals that uptake of a complete course of chemotherapy results in better treatment outcomes. Among the SIII, NLR, and PNLR are of better prognostic value and could be used as alternative tools for predicting treatment outcomes.

## Data Availability

All data produced in the present work are contained in the manuscript

## References

1. Ferlay J, Colombet M, Soerjomataram I, Parkin DM, Piñeros M, Znaor A, Bray F. Cancer statistics for the year 2020: An overview. Int J cancer. 2021;149(4):778–89.

2. Chornokur G, Amankwah EK, Schildkraut JM, Phelan CM. Global ovarian cancer health disparities. Gynecol Oncol. 2013;129(1):258–64.

3. Gizaw M, Parkin DM, Stöter O, Korir A, Kamate B, Liu B, Bojang L, N’Da G, Manraj SS, Bukirwa P, Chokunonga E. Trends in the incidence of ovarian cancer in sub-Saharan Africa. Intl J Cancer. 2023; 152(7):1328–36.

4. Bray F, Ferlay J, Soerjomataram I, Siegel RL, Torre LA, Jemal A. GLOBOCAN estimates of incidence and mortality worldwide for 36 cancers in 185 countries: Global cancer statistics. CA Cancer J Clin. 2018;68(6):394–424.

5. Torre LA, Trabert B, DeSantis CE, Miller KD, Samimi G, Runowicz CD, Gaudet MM, Jemal A, Siegel RL. Ovarian cancer statistics, 2018. CA: a cancer J Clin. 2018; 68(4):284–96.

6. Hirose S, Tanabe H, Nagayoshi Y, Hirata Y, Narui C, Ochiai K, Isonishi S, Takano H, Okamoto A. Retrospective analysis of sites of recurrence in stage I epithelial ovarian cancer. Journal of Gynecol Oncol. 2018;29(3).

7. Allensworth SK, Langstraat CL, Martin JR, Lemens MA, McGree ME, Weaver AL, Dowdy SC, Podratz KC, Bakkum-Gamez JN. Evaluating the prognostic significance of preoperative thrombocytosis in epithelial ovarian cancer. Gynecol Oncol. 2013;130(3):499–504.

8. Webb PM, Jordan SJ. Epidemiology of epithelial ovarian cancer. Best Prac Res Clin Obstet gynaecol 2017;41:3–14.

9. Kotsopoulos J, Gronwald J, Karlan B, Rosen B, Huzarski T, Moller P, Lynch HT, Singer CF, Senter L, Neuhausen SL, Tung N. Age-specific ovarian cancer risks among women with a BRCA1 or BRCA2 mutation. Gynecologic oncology. 2018;150(1):85–91.

10. Bizzarri N, D’Indinosante M, Marchetti C, Tudisco R, Turchiano F, Scambia G, Fagotti A. The prognostic role of systemic inflammatory markers in apparent early-stage ovarian cancer. Int J Clin Oncol. 2023;28(2):314–20.

11. Marchetti C, D’Indinosante M, Bottoni C, Di Ilio C, Di Berardino S, Costantini B, Minucci A, Vertechy L, Scambia G, Fagotti A. NLR and BRCA mutational status in patients with high grade serous advanced ovarian cancer. Sci Rep. 2021;11(1):11125.

12. Huang J, Hu W, Sood AK. Prognostic biomarkers in ovarian cancer. Cancer Biomarkers. 2011; 8(4-5):231–51.

13. Muinao T, Deka Boruah HP, Pal M. Diagnostic and Prognostic Biomarkers in ovarian cancer and the potential roles of cancer stem cells—an updated review. Exp Cell Res 362(1):1–10.

14. Grivennikov SI, Greten FR, Karin M (2010) Immunity, inflammation, and cancer. Cell 140(6):883–899. https://doi.org/10.1016/j.cell.2010.01.025

15. Proctor MJ, McMillan DC, Horgan PG, Fletcher CD, Talwar D, Morrison DS. Systemic inflammation predicts all-cause mortality: a Glasgow inflammation outcome study. PloS one. 2015;10(3):e0116206.

16. Templeton AJ, McNamara MG, Šeruga B, et al (2014) Prognostic role of neutrophil-to-lymphocyte ratio in solid tumors: a systematic review and meta-analysis.

17. Templeton AJ, McNamara MG. B. eruga, FE Vera-Badillo, P. Aneja, A. Ocaa, R. Leibowitz-Amit, G. Sonpavde, JJ Knox, B. Tran, IF Tannock, and E. Amir,”Prognostic role of neutrophil-to-lymphocyte ratio in solid tumors: A systematic review and meta-analysis,” J Natl Cancer Inst 106(6):dju124.

18. Dumitru CA, Lang S, Brandau S. Modulation of neutrophil granulocytes in the tumor microenvironment: Mechanisms and consequences for tumor progression Semin Cancer Biol. 2013;23:141–8

19. Zhu Y, Zhou S, Liu Y, Zhai L, Sun X. Prognostic value of systemic inflammatory markers in ovarian Cancer: a PRISMA-compliant meta-analysis and systematic review. BMC cancer. 2018; 18(1):1–0.

20. El Bairi K, Al Jarroudi O, Afqir S. Inexpensive systemic inflammatory biomarkers in ovarian Cancer: an umbrella systematic review of 17 prognostic meta-analyses. Frontiers Oncol. 2021;11:694821.

21. Huang H, Wu K, Chen L, Lin X. Study on the application of systemic inflammation response index and platelet–lymphocyte ratio in ovarian malignant tumors. Intl J General Med. 2021:10015–22.

22. Mleko M, Pluta E, Pitynski K, Bodzek M, Kałamacki A, Kiprian D, Banas T. Trends in Systemic Inflammatory Reaction (SIR) during Paclitaxel and Carboplatin Chemotherapy in Women Suffering from Epithelial Ovarian Cancer. Cancers. 2023;15(14):3607.

23. Iyoke CA, Ugwu GO, Ezugwu EC, Onah N, Ugwu O, Okafor O. Incidence, pattern and management of ovarian cancer at a tertiary medical center in Enugu, Southeast Nigeria. Ann Med Health Sci Res. 2013 Sep 18;3(3):417–21.

24. Okunade KS, Okunola H, Okunowo AA, Anorlu RI. A five-year review of ovarian cancer at a tertiary institution in Lagos, South West, Nigeria. Accessed on 18/07/23. Available at https://ir.unilag.edu.ng/handle/123456789/10071

25. Zayyan MS, Ahmed SA, Oguntayo AO, Kolawole AO, Olasinde TA. Epidemiology of ovarian cancers in Zaria, Northern Nigeria: a 10-year study. Int J Women’s Health. 2017 Nov 22:855–60.

26. Okunade KS, Adejimi AA, Ohazurike EO, Salako O, Osunwusi B, Adenekan MA, Ugwu AO, Soibi-Harry A, Dawodu O, Okunowo AA, Anorlu RI. Predictors of survival outcomes after primary treatment of epithelial ovarian cancer in Lagos, Nigeria. JCO Global Oncol. 2021;7:89–98.

27. Lee JY, Jeon I, Kim JW, Song YS, Yoon JM, Park SM. Diabetes mellitus and ovarian cancer risk: a systematic review and meta-analysis of observational studies. Int J Gynecol Cancer. 2013;23(3).

28. Li Y, Jia H, Yu W, Xu Y, Li X, Li Q, Cai S. Nomograms for predicting the prognostic value of inflammatory biomarkers in colorectal cancer patients after radical resection. Intl J Cancer. 2016; 139(1):220–31.

29. Gong TT, Wu QJ, Vogtmann E, Lin B, Wang YL. Age at menarche and risk of ovarian cancer: a meta-analysis of epidemiological studies. Int J cancer. 2013;132(12):2894–900.

30. Alegbeleye BJ, Akpoveso OO, Mohammed RK. The use of herbal medicines by cancer patients in contemporary African settings: a scoping review. Int J Sci Adv. 2020;1:49–73.

31. Zhu K, Fukasawa I, Furuno M, Inaba F, Yamazaki T, Kamemori T, Kousaka N, Ota Y, Hayashi M, Maehama T, Inaba N. Inhibitory effects of herbal drugs on the growth of human ovarian cancer cell lines through the induction of apoptosis. Gynecol Oncol; 97(2):405–9.

32. Ben-Arye E, Lavie O, Samuels N, Khamaisie H, Schiff E, Raz OG, Mahajna J. Safety of herbal medicine use during chemotherapy in patients with ovarian cancer: a “bedside-to-bench” approach. Med Oncol. 2017 Apr;34:1–6.

33. Awodele O, Popoola TD, Amadi KC, Coker HA, Akintonwa A. Traditional medicinal plants in Nigeria—Remedies or risks. J Ethnopharmacol. 2013; 150(2):614–8.

34. Mrozek-Szetela A, Rejda P, Wińska K. A review of hygienization methods of herbal raw materials. Appl Sci. 2020;10(22):8268.

35. Başaran N, Paslı D, Başaran AA. Unpredictable adverse effects of herbal products. Food Chem Toxicol. 2022;159:112762.

